# An Analysis of Self-reported Longcovid Symptoms on Twitter

**DOI:** 10.1101/2020.08.14.20175059

**Authors:** Shubh Mohan Singh, Chaitanya Reddy

**Affiliations:** Postgraduate Institute of Medical Education and Research Chandigarh India

**Keywords:** Longcovid, longhauler, COVID-19, persistent symptoms, symptomatology, twitter, social media

## Abstract

**Objectives:** A majority of patients suffering from acute COVID-19 are expected to recover symptomatically and functionally. However there are reports that some people continue to experience symptoms even beyond the stage of acute infection. This phenomenon has been called longcovid.

**Study design:** This study attempted to analyse symptoms reported by users on twitter self-identifying as longcovid.

**Methods:** The search was carried out using the twitter public streaming application programming interface using a relevant search term.

**Results:** We could identify 89 users with usable data in the tweets posted by them. A majority of users described multiple symptoms the most common of which were fatigue, shortness of breath, pain and brainfog/concentration difficulties. The most common course of symptoms was episodic.

**Conclusions:** Given the public health importance of this issue, the study suggests that there is a need to better study post acute-COVID symptoms.

---

The COVID-19 pandemic has affected millions of people across the world. It is expected that a majority of the patients with COVID-19 would recover from infection ^1^. However there are reports that many of these people continue to experience symptoms and disability ^2^. In english speaking countries, the term “longcovid” or “longahulers” has been used to describe this phenomenon and the people suffering from it respectively ^3,4^. There are various anecdotal reports, discussions on online forums and social media, and patient led research regarding the experiences of people who self-identify as suffering from longcovid ^5^. Social media such as twitter has been used to mine data regarding patient accounts of symptomatology in the COVID-19 pandemic ^6^. Given the public health importance of this issue and paucity of information from conventional sources, this study was undertaken to analyse symptoms associated with longcovid reported by users on twitter.

Tweets were collected using the rtweet package in RStudio software from the twitter public streaming application programming interface. Given the millions of tweets on COVID-19, the search protocol was restricted to the term “longcovid”, tweets in english language, and retweets were excluded. The retry on rate limit function was set to true to retrieve as many tweets as possible. Once the tweets were retrieved, these were then manually read. Tweets that were not in english, or by users who did not identify themselves as having longcovid symptoms or having symptoms due to another disorder such as lyme disease or chronic fatigue syndrome/myalgic encephalomyelitis, tweets about longcovid in general but not experienced symptoms excluded. The tweets included in final analysis were in english by users who identified as having longcovid and the content of the tweets pertained to the symptoms experienced by them.

The initial search retrieved 3449 tweets from 190 different tweeters from 20th July 2020 to 29th July 2020. After manually reading the tweets and applying the inclusion criteria, 165 tweets from 89 users were included in the final analysis. Multiple tweets from single users were collapsed into one text block and repeat or similar symptoms were deleted so that symptoms referred to in more than one tweet were counted only once per person. 240 symptoms were mentioned by all the users. On analysis, these were grouped into 94 different symptoms. The most common symptoms with > 1 mentions are presented in table 1. 47 users mentioned the time period of symptoms and these ranged from 3 weeks to 42 weeks (16–20 weeks by 32 users). There was no association between nature and number of symptoms reported and the duration of illness. 29 users mentioned the course of their symptoms. The most pattern described was one of episodes or relapses (n=16), followed by a continuous course (n=9), of which some described fluctuations in the course of symptoms (n=3) and 4 users described continuous symptoms with added on symptoms during exacerbations. The common precipitating factors for exacerbations were physical activity (n=3), trauma (n=1) and heat (n=1). 53 users (59.55%) reported more than one symptom. Unique hashtags associated with longcovid included ‘longhaulers’ and ‘NEISVOID’.

**Table 1.** Symptoms with > 1 mention in tweets

| Order | Symptoms | Prevalence (%) |
| --- | --- | --- |
| 1. | Fatigue | 42 (47.19) |
| 2. | Shortness of breath | 23 (25.84) |
| 3. | Brainfog | 15 (16.85) |
| 4. | Exercise intolerance | 13 (14.60) |
| 5. | Pain in the whole body | 9 (10.11) |
| 6. | Altered smell | 7 (7.86) |
| 7. | Headache | 7 (7.86) |
| 8. | Tachycardia | 6 (6.74) |
| 9. | Altered taste | 6 (6.74) |
| 10. | Pain chest | 5 (5.61) |
| 11. | Dizziness | 3 (3.37) |
| 12. | Pain abdomen | 3 (3.37) |
| 13. | Fever | 3 (3.37) |
| 14. | Nausea | 3 (3.37) |
| 15. | Cough | 3 (3.37) |
| 16. | Weakness | 3 (3.37) |
| 17. | Altered appetite | 2 (2.24) |
| 18. | Pain (location not specified) | 2 (2.24) |
| 19. | Chest tightness | 2 (2.24) |
| 20. | Numbness | 2 (2.24) |
| 21. | Covid symptoms | 2 (2.24) |
| 22. | Tinnitus | 2 (2.24) |
| 23. | Altered sleep | 2 (2.24) |
| 24. | Pain in ear | 2 (2.24) |
| 25. | Pain in muscles | 2 (2.24) |
| 26. | Irritability | 2 (2.24) |
| 27. | Tachycardia on exertion | 2 (2.24) |

Twitter data mining has been used as a means for analysis of symptom profiles in various disorders including COVID-19 ^6^. There are various limitations in non-experimental twitter derived data including patchy incomplete data, the self selection of users, lack of objective validations of reported symptoms, inexact reporting of variables such as sociodemographic data and non-exact definitions of symptomatology ^7^. There is also the possibility of relevant data being missed or being available on other social media platforms. Our data set also suffers from some of these limitations. In addition we did not have any information regarding the symptom severity of initial disease and treatment details.

There are anecdotal reports, online content and some evidence to suggest that a proportion of patients who have recovered from acute COVID-19 continue to have some symptoms ^2,8^. Considering the millions of people who have been infected and recovered from acute COVID-19, persistent symptoms and presumably disability and impairment in quality of life may pose a significant public health challenge. The results of this analysis shows that the signal to noise ratio with regards to patient driven data in COVID-19 is very high as has been observed earlier ^6^. This may reflect the interest generated by COVID-19, also we observed that longcovid related content also attracted users identifying with the lyme disease, myalgic encephalomyelitis/chronic fatigue syndrome, fibromyalgia communities who offered support and identified symptoms reported as longcovid to be similar to theirs.

In keeping with the limited data available, this analysis shows that a majority of the users reported multiple symptoms (>1) at varying durations of illness (3 weeks to 42 weeks with a majority at 16–20 weeks). This was similar to that reported in another study even though the average duration of post acute assessment was at 60 days (∼9weeks)^2^. The most common symptoms reported were also similar (fatigue, shortness of breath, brainfog/concentration difficulties, pain, cough, disturbances in smell, taste and appetite, chest discomfort, headache etc.). In addition, users described different courses of symptoms with most describing an episodic or relapsing course usually brought about by some precipitant.

Chronic persistence of symptoms in post acute-SARS infection has been described earlier with somewhat symptomatologies ^9^. Some have speculated that these could be due to immunological mechanisms ^8^. Similarly, post viral fatigue syndromes have been described due to a variety of etiologies ^10^. These are often associated with chronicity and development of chronic fatigue syndrome ^10^. The result of this twitter analysis suggests that some patients who have recovered from acute COVID-19 seem to have multiple prolonged symptoms (as described above) with varying courses and outcomes.

There is a need to better characterise post COVID-19 symptomatology and elucidate the mediating variables through rigorous scientific studies. If present in significant numbers of people, the prevention and management of these symptoms should be incorporated in treatment protocols.

## Data Availability

Data is available

## Acknowledgments

None

This research did not receive any specific grant from funding agencies in the public, commercial, or not-for-profit sectors.

Conflict of interest

None

